# Architecture of systems affecting disease trajectories in a conflict zone: A community-centered systems inquiry in North Gaza

**DOI:** 10.1101/2025.03.11.25323741

**Authors:** Yahya Shaikh, Mohammed O. A. Hamouda, Shatha Elnakib

## Abstract

Humanitarian crises, particularly in conflict zones, create cascading disruptions that impact every aspect of daily life, including health and disease outcomes. While international humanitarian frameworks categorize these crises into discrete operational clusters, affected populations experience them as interwoven, systemic failures. This study examines how conflict-induced disruptions transform a preventable and typically self-limiting disease— Hepatitis A—into a fatal outcome in North Gaza. Using a systems approach, we seek to characterize the architecture of interconnected disruptions leading to preventable deaths.

This study employed the FAIR (Fairness, Agency, Inclusion, and Representation) Framework, a participatory methodology centering community epistemes, to analyze four pediatric cases of Hepatitis A that progressed to fulminant liver failure. Data were obtained through interviews with healthcare providers, caregivers, and community members, supplemented by medical chart reviews. A network-based Architecture of Systems (AoS) map was constructed to visualize the interconnections between war-induced systemic disruptions and health outcomes. Network analysis was performed to identify key nodes, bottlenecks, and pathways within this system.

The findings of this study reveal a complex system of war-driven factors—displacement, destruction of healthcare infrastructure, water scarcity, food deprivation, and fuel blockades— that collectively reshaped disease trajectories. Network analysis of the AoS map identified 138 nodes and 231 edges, generating 41,444 pathways linking conflict-related disruptions to health outcomes. Women’s health emerged as a central mediator, with 95% of pathways intersecting with 13 key nodes related to women’s roles in caregiving, resource acquisition, and psychological stability. The lack of access to food and clean water, combined with the destruction of healthcare facilities and restrictions on medical evacuation, created conditions where a preventable disease became fatal.

This study highlights how conflict restructures health determinants, shaping survival strategies that paradoxically increase morbidity and mortality. By documenting the lived experiences of affected communities, our findings underscore the necessity of a systems-based humanitarian response that accounts for the interconnected nature of crises. Recognizing the pivotal role of women in mediating health outcomes is crucial to designing effective interventions in conflict settings.

## Introduction

Humanitarian crises, particularly in times of conflict, are rarely experienced by affected populations as isolated events or discrete challenges. Instead, they manifest as interlinked, cascading systems of disruption that reshape every aspect of daily life, force movement, restrict choices, dictate behaviors, and ultimately worsen health, disease, and mortality. (1–3) While the humanitarian sector often deconstructs these crises into operational clusters—such as food security, healthcare, or shelter—to facilitate aid,(4–6) this fragmented perspective risks overlooking the lived experiences of those enduring the crisis.(3,7) For affected populations, these sectors are inseparable components of a deteriorating or adversely reconstructed system. The failure to represent this interconnected system means that representations of the affected community’s realities are incongruent to their lived experiences.(8–14) This, in effect, results in silencing the community’s voice, (15) upholding a system that has adverse impacts on the population, (11,16–18) and constitutes epistemic violence.

Epistemic violence occurs when the experience of affected populations is reframed by external actors in ways that distort or diminish their reality.(15,19–23) In humanitarian crises, this is particularly problematic, as aid frameworks designed for logistical clarity often fail to capture the systemic interdependence of events as they are experienced by the community.(7) For instance, the bombing of residential areas may lead to displacement, which, coupled with restrictions on food and water transport, creates conditions for malnutrition and disease outbreaks. These interconnected phenomena are not experienced as separate “sectors” but as a coherent, worsening system of survival challenges.

Emerging research emphasizes the need for a systems perspective in understanding and responding to humanitarian crises.(3) By examining humanitarian crises as systems of effects, researchers and practitioners can more accurately represent the complex realities of affected populations, ensuring that interventions address not only individual factors but also the relationships between them.(7,24) This perspective is essential to ensure that a population attempting to survive a humanitarian crises is not additionally violated by epistemic violence in the name of humanitarian relief.(23)

Hepatitis A provides an important lens for applying this systems perspective because it illustrates how interconnected systemic failures during humanitarian crises can transform a preventable, self-limiting disease into a fatal outcome. Studies have shown that conflict-induced conditions—such as overcrowding, lack of sanitation, and destruction of water systems—create environments where fecal-oral transmission diseases like Hepatitis A spread rapidly.(25,26) Simultaneously, restricted access to healthcare, exacerbated by destroyed infrastructure and medical supply shortages, limits timely diagnosis and treatment, allowing complications to escalate.(27) These interacting factors—arising from systemic disruptions—highlight the need to view health outcomes not as isolated clinical phenomena but as products of broader socio-political and infrastructural destruction.

In this article, we characterize the system leading to child deaths from Hepatitis A, a rare complication of a preventable disease.(28) Using the FAIR (Fairness, Agency, Inclusion, and Representation) Framework,(29) a participatory research methodology to center community epistemes, we ask and attempt to answer the question: What is the architecture of the system of interconnected disruptions during a humanitarian crisis that transforms a preventable disease (like Hepatitis A) into a fatal outcome?

This inquiry seeks to uncover the interconnected pathways through which socio-political and infrastructural destruction amplify risks, restrict choices, and shape survival strategies, ultimately contributing to adverse health outcomes. By centering the lived experiences of the affected community, this study attempts to uncover mechanisms of morbidity and mortality when civilian populations are impacted by military operations.

## Methods

### Study setting and design

This study was conducted in the period spanning June 27^th^, 2024 to July 25^th^, 2024 in North Gaza during an extreme humanitarian crisis amid an ongoing military campaign by Israel that began in October 2023. Since the war’s escalation, North Gaza has been under a “siege within a siege,” with severe restrictions on food and medical supplies. By March-April 2024, these restrictions resulted in widespread hunger and malnutrition, with the civilian population reaching Phase 5 (catastrophic hunger) of the Integrated Food Security Phase Classification. Many residents survived by consuming animal feed and foraged weeds.

This study aimed to examine the systemic factors contributing to deaths from Hepatitis A, a typically preventable disease, by focusing on four pediatric patient cases who presented to hospitals in North Gaza between March and April 2024. The study used a participatory methodology to engage healthcare providers, families, and caregivers to uncover systemic barriers that contributed to these fatalities.

### Participants

The study focused on four pediatric patients who experienced disease progression to fulminant liver failure and death. Participants included in the study were as follows:

- Healthcare providers involved in the care of these four patients.
- Family members and caregivers who provided additional insights into patient health journeys.
- Community members familiar with the broader socio-political context shaping access to healthcare.

Given the extreme security risks and constraints, participant engagement was adapted to ensure feasibility and safety, including condensed sessions, flexible meeting times, and modified data collection methods.

### Data Collection and Analysis

The study employed the FAIR Framework, a participatory approach to knowledge production that prioritizes the perspectives and epistemologies of communities, recognizing the agency of communities in representing and interpreting their own experiences.(23,29) It engages community members to convey the context of their lived experience and to translate their insights into systems of understanding that inform external researchers and policymakers. This approach bridges the gap between community epistemologies and dominant systems of knowledge, creating outputs that are more accurate, equitable, and aligned with the lived experiences of those on the margins.(29)

The FAIR Framework consists of a multi-step process designed to center community knowledge and experiences in understanding systemic challenges. It begins with Knowledge-Bearer and Knowledge-Interpreter recruitment, identifying community members with deep, lived expertise and the ability to articulate and contextualize their community’s realities. This is followed by relationship building to establish trust, ensure mutual understanding, and create a collaborative space for exploring community knowledge. Next, in vignette development and enrichment, participants craft detailed narratives that reflect diverse lived experiences within the community, which are then analyzed to identify interconnected systemic factors. These analyses led to the construction of an architecture of systems map, a visual representation of the relationships between key factors shaping systemic challenges. The map is subsequently augmented using literature and additional insights to enhance its accuracy and applicability. Each step is iterative and participatory, ensuring that outputs authentically represent the community’s epistemologies while fostering actionable insights for addressing systemic inequities.

The methodology was adapted to the war context in Gaza and structured into three phases:

1. Patient Clinical Journey and Medical Chart Procurement:

◦ Reviewed patient charts of pediatric patients who had hepatitis A, developed fulminant hepatic failure and died from its sequelae between March 1^st^, 2024 and April 30^th^, 2024.
◦ Integrated clinical history elicitation within the patient record process.
2. Socio-Political Context and Architecture of Systems Mapping:

◦ Engaged caregivers, providers, and community members to construct a contextualized patient health journey.
◦ Developed an Architecture of Systems Map to represent systemic barriers.
◦ Used iterative coding and thematic clustering to refine cause-effect relationships.
3. Community Validation of the Systems Map:

◦ Healthcare providers and caregivers validated the constructed systems map, ensuring it accurately represented patient experiences.

Each of the above steps are guided by participatory modeling scripts (S1 File). Once the architecture of systems diagram was constructed, network analysis was conducted to identify key nodes and their centrality measures. The analysis aimed to uncover the most connected, influential, and accessible nodes in the network. Key network metrics were calculated to understand the structure and significance of nodes within the network and included:

- Degree centrality: the number of direct connections a node has (both incoming and outgoing), normalized by the maximum possible connections in the network. This metric identified the most connected nodes, representing highly influential factors or those most affected
- Betweenness centrality: the frequency with which a node acts as a bridge along the shortest paths between other nodes. Nodes with high betweenness centrality indicate critical bottlenecks or intermediaries where interventions may have the greatest cascading effects
- Closeness centrality (factors most accessible within the network): the closeness a node is to all other nodes in the network, measured as the reciprocal of the average shortest path length from the node to all others. Nodes with high closeness centrality are centrally positioned and readily accessible, indicating factors that can rapidly influence or be influenced by others

Network analysis was performed using Python version 3.11.

### Ethics Approval

The study protocol was reviewed by Johns Hopkins University IRB (IRB No: 31883) and received an exemption as it involved the use of existing secondary data. To ensure confidentiality and security, no individual identifiers were recorded. Verbal informed consent was obtained from all participants prior to participation, further safeguarding privacy amid security risks. Discussion groups were conducted by the first two authors.

## Results

This study included 25 participants (7 healthcare workers; 18 community members including parents, family, and neighbors) with a mean age of 35+/-9 years, of whom 10 (40%) were women. Using the FAIR Framework, over a span of on average 2 sessions per person, with some sessions occurring with multiple people at the same time for a total of 19 sessions, the study explored the system that shapes patient trajectories of a preventable and self-limited disease towards fatal outcomes in North Gaza. The findings reveal a complex interplay of precipitating factors and pathways driven by war-induced disruptions, highlighting the systemic nature of the humanitarian crisis.

The patient journeys obtained from medical charts, providers, and caregivers (Phase 1 of the modified FAIR Framework) related to four otherwise healthy children with similar histories and clinical trajectories who presented with hepatitis A which subsequently progressed to fulminant hepatic failure and death. All four children were previously healthy, aged 7-10 years old, brought at various times between April and June 2024 to the Emergency Department of a community hospital in North Gaza after several weeks of progressively worsening symptoms that included abdominal pain, loss of appetite, nausea, vomiting, diarrhea, icterus, and jaundice. They had initial presentations to the healthcare system where they were appropriately managed conservatively for malnutrition and hepatitis A infection. After several subsequent weeks of worsening symptoms, they were finally brought in due to lethargy and changing levels of consciousness. Their social histories were significant for multiple displacements from Internally Displaced Persons (IDP) camps due to evacuation orders, after their homes were initially destroyed in air strikes by Israeli forces. They all experienced a period of severe hunger from January 2024 to April 2024, during which time they consumed animal feed, often ground and incorporated as part of the “flour” for bread. Their risk factors (i.e. poor sanitation, limited access to clean water, crowded conditions), symptoms, clinical findings, lab tests, and imaging were indicative of acute malnutrition and hepatitis A infection with hepatic encephalopathy. The siege and ongoing air strikes on civilian infrastructure restricted the ability to diagnose and monitor, treatment options, referral options, transport to neighboring facilities for higher levels of care for all patients.

Centering community voice to describe factors that shaped patient trajectories (Phase 2 of the modified FAIR Framework) led to the generation of 138 codes whose relationships were visualized in the AoS diagram (Fig 1). In the AoS diagram, precipitating or ultimate causal factors were thematically identified and organized in clusters along the X-axis, while cause-effect relationships were thematically identified and separated along the Y-axis.

**Fig 1.**
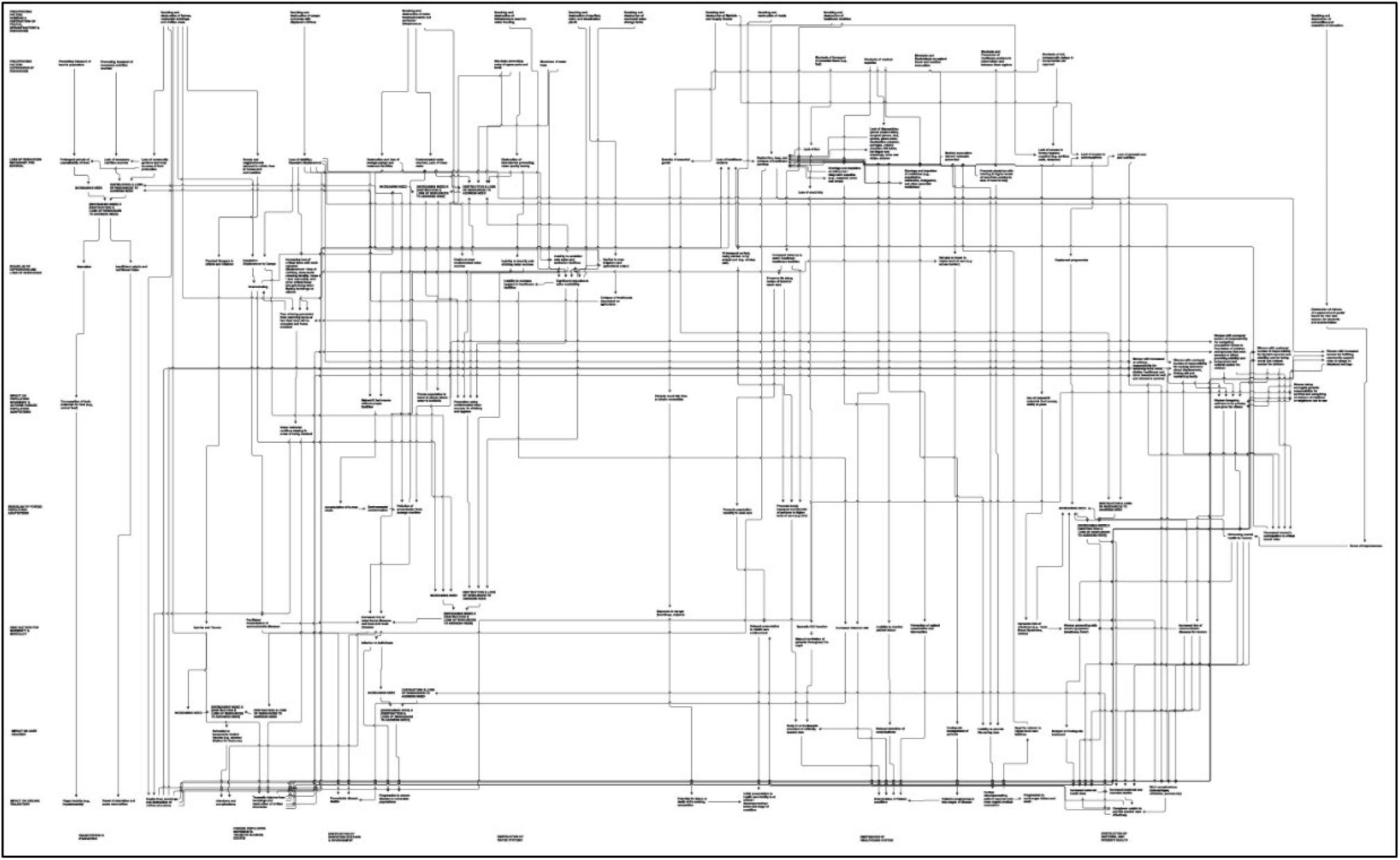
Architecture of Systems Diagram.

Grouping related codes together resulted in 12 thematic categories of effects (Table 1), some of which include civilian deaths, starvation, maternal and neonatal morbidity and mortality, multiple displacements, loss of visions hope; in addition to destruction of community systems of food, water, and health care. The cause-effect relationships between codes formed 11 categories of relationships (Fig 2), which included: bombing and destruction along with deprivation of resources lead to impacts on the population increasing need for health care (loss of resources necessary for survival; sequelae of deprivation and loss of resources; population adaptations; consequences of those adaptations, and increased risks for morbidity and mortality), while at the same time reducing resources and ability to provide health care.

**Fig 2.**
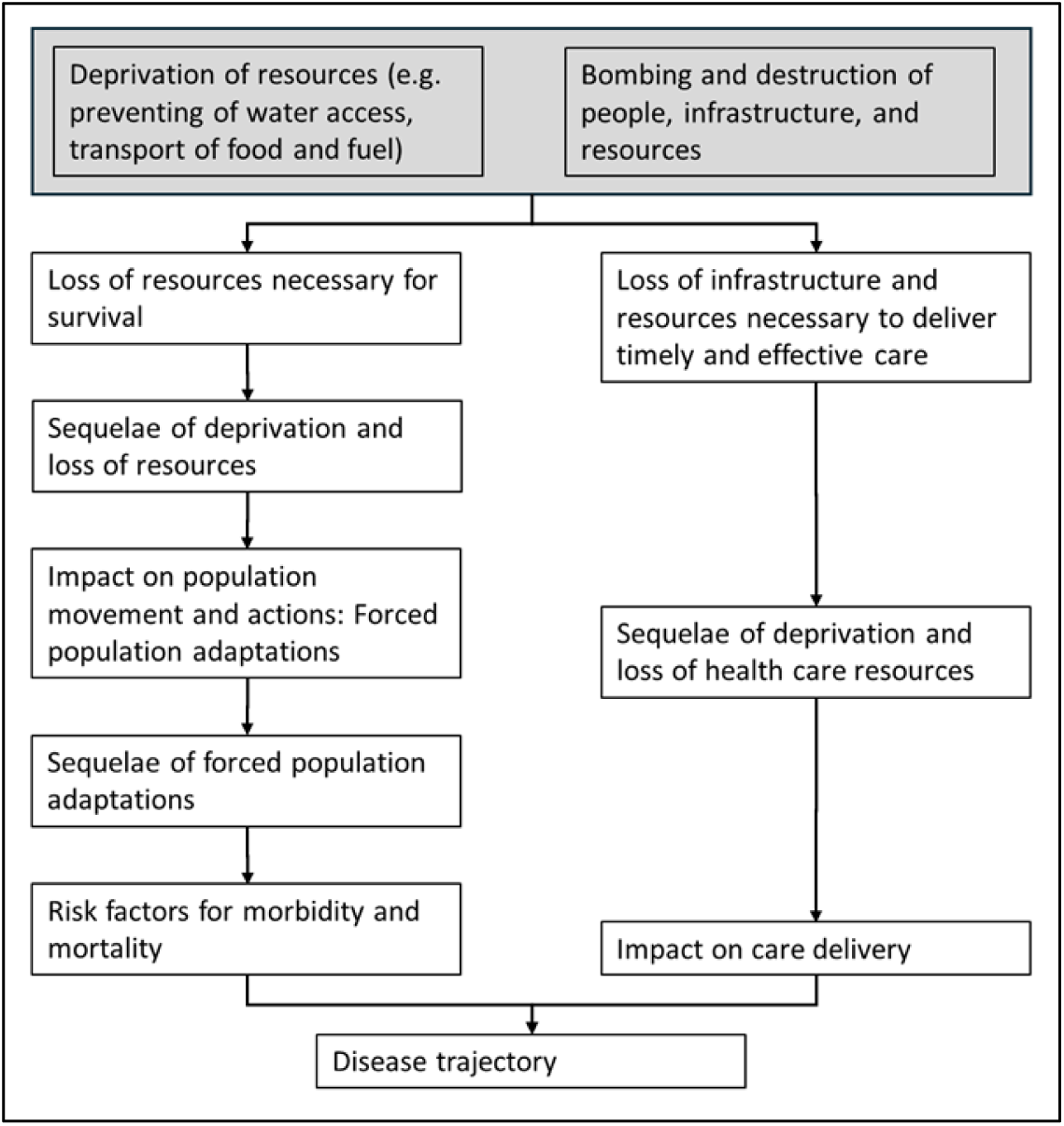
Thematic Categories of Cause-Effect Relationships.

**Table 1.** Thematic categories of primary codes.

| High-Level Categories | Thematic Categories of Primary Codes |
| --- | --- |
| Effect of bombing and deprivation on people | Starvation of population |
|  | Deaths; traumatic injuries |
|  | Maternal and neonatal morbidity and mortality |
|  | Destruction of homes and shelter |
|  | Loss of personal items necessary for normal life: documents and certificates, identification papers, valuables; books, medicines, clothing, household items |
|  | Forced population movements; multiple displacements |
|  | Loss of hope and visions of the future |
| **Effect of bombing and deprivation on population resources** | Destruction of food systems |
|  | Destruction of water systems |
|  | Destruction of healthcare system |
|  | Destruction of education system |
|  | Destruction of sanitation systems; contamination of environment |

The results of the network analysis show a graph with 138 nodes and 231 edges, resulting in 41,444 pathways. Notably, 13 nodes (9%) related to women’s health while 39,639 (95%) pathways connected to women’s health through these nodes (see S1 FIle1, Table A; and S2 File). The top 10 nodes by centrality are given in Table 2. The most connected nodes (i.e. degree centrality) were: Destruction, loss, and collapse of healthcare services; Loss of stability; Repeated displacement; Significant reduction in water availability; Traumatic injuries and deaths from bombings and destruction of civilian structures; and the impact of the war on women’s physical and mental health.

**Table 2.**
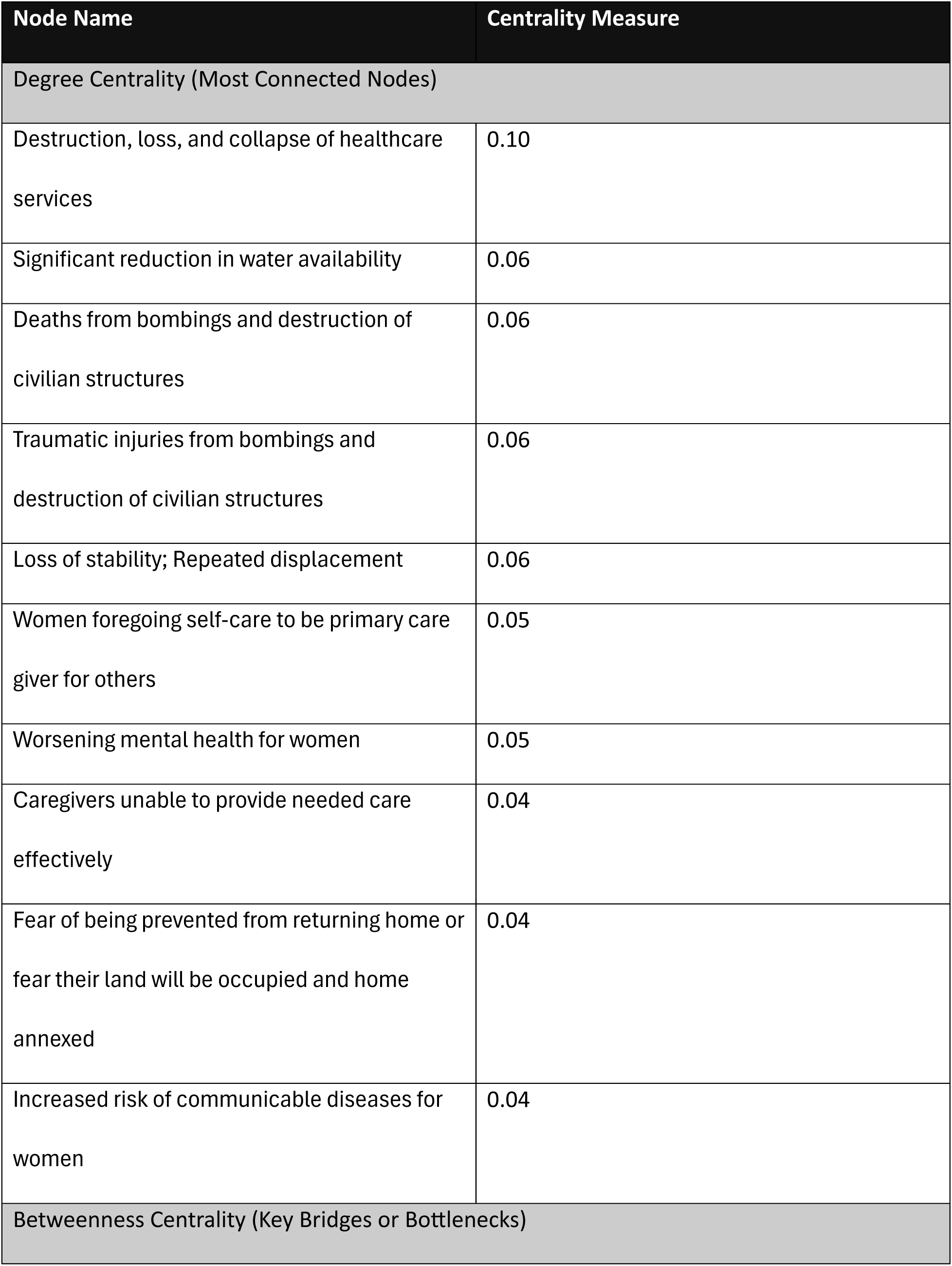
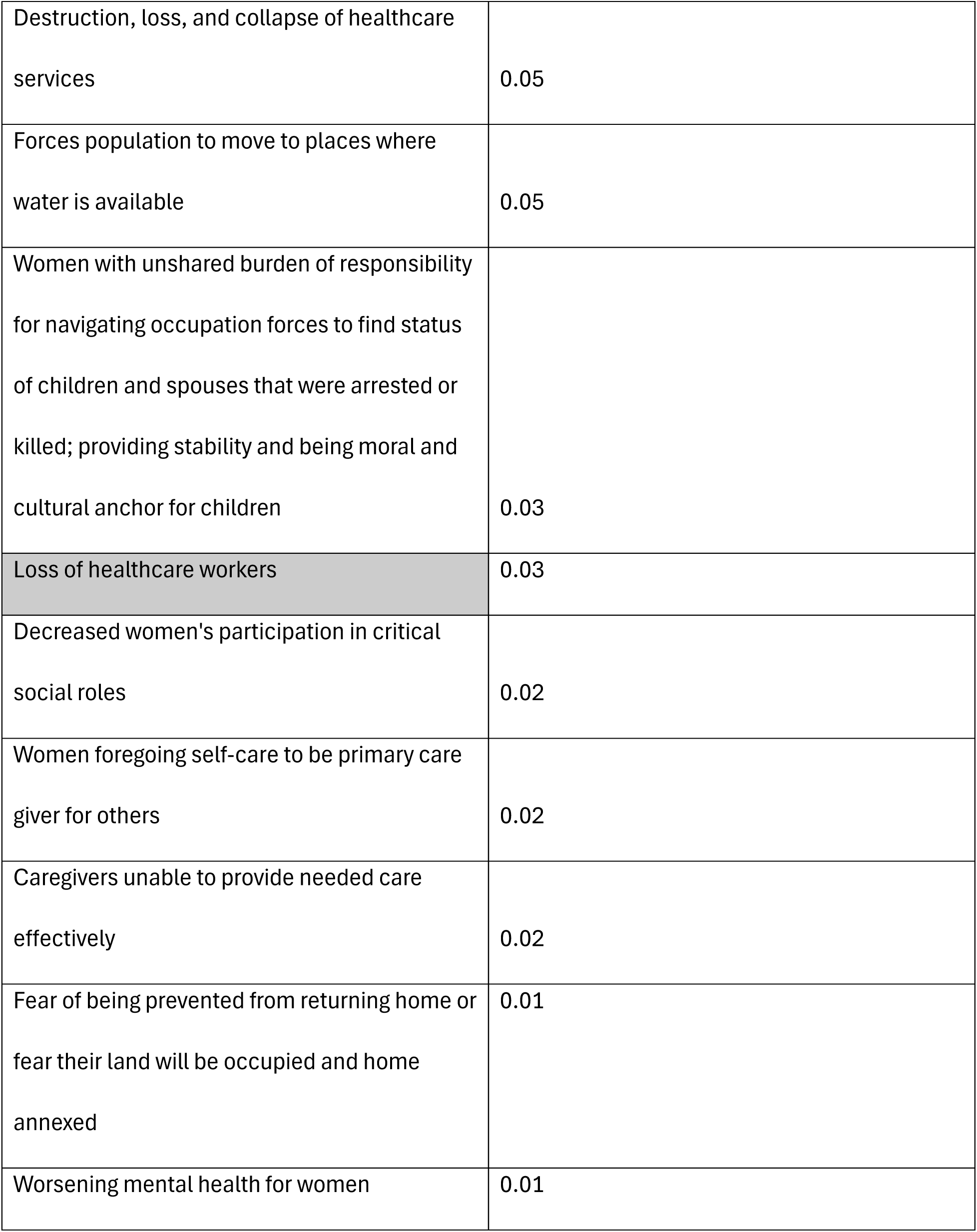
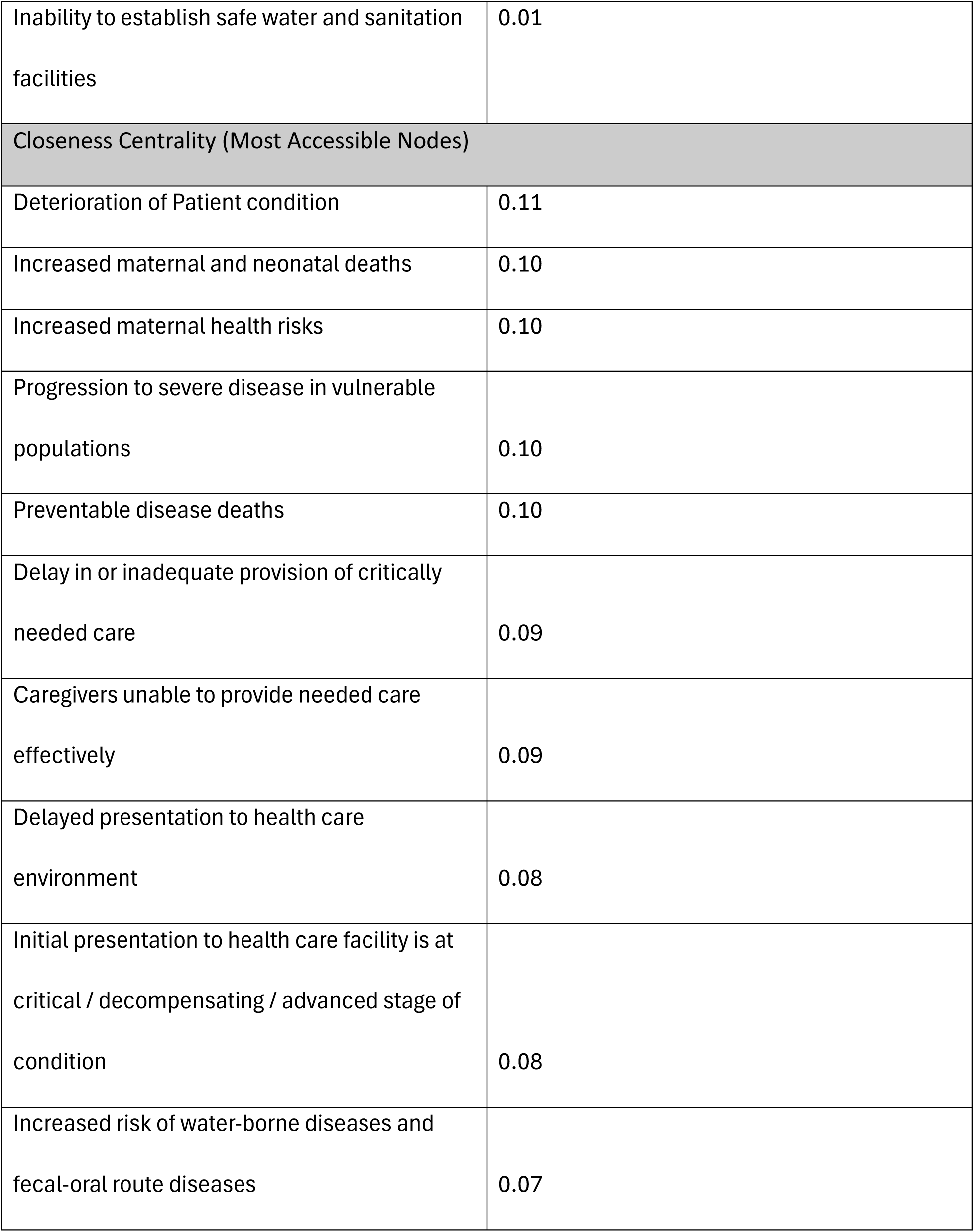
Centrality Measures of Architecture of Systems Map.

| Node Name | Centrality Measure |
| --- | --- |

| Degree Centrality (Most Connected Nodes) |  |
| --- | --- |
| Destruction, loss, and collapse of healthcare services | 0.10 |
| Significant reduction in water availability | 0.06 |
| Deaths from bombings and destruction of civilian structures | 0.06 |
| Traumatic injuries from bombings and destruction of civilian structures | 0.06 |
| Loss of stability; Repeated displacement | 0.06 |
| Women foregoing self-care to be primary care giver for others | 0.05 |
| Worsening mental health for women | 0.05 |
| Caregivers unable to provide needed care effectively | 0.04 |
| Fear of being prevented from returning home or fear their land will be occupied and home annexed | 0.04 |
| Increased risk of communicable diseases for women | 0.04 |
| Betweenness Centrality (Key Bridges or Bottlenecks) |  |
| Destruction, loss, and collapse of healthcare services | 0.05 |
| Forces population to move to places where water is available | 0.05 |
| Women with unshared burden of responsibility for navigating occupation forces to find status of children and spouses that were arrested or killed; providing stability and being moral and cultural anchor for children | 0.03 |
| Loss of healthcare workers | 0.03 |
| Decreased women's participation in critical social roles | 0.02 |
| Women foregoing self-care to be primary care giver for others | 0.02 |
| Caregivers unable to provide needed care effectively | 0.02 |
| Fear of being prevented from returning home or fear their land will be occupied and home annexed | 0.01 |
| Worsening mental health for women | 0.01 |
| Inability to establish safe water and sanitation facilities | 0.01 |
| Closeness Centrality (Most Accessible Nodes) |  |
| Deterioration of Patient condition | 0.11 |
| Increased maternal and neonatal deaths | 0.10 |
| Increased maternal health risks | 0.10 |
| Progression to severe disease in vulnerable populations | 0.10 |
| Preventable disease deaths | 0.10 |
| Delay in or inadequate provision of critically needed care | 0.09 |
| Caregivers unable to provide needed care effectively | 0.09 |
| Delayed presentation to health care environment | 0.08 |
| Initial presentation to health care facility is at critical / decompensating / advanced stage of condition | 0.08 |
| Increased risk of water-borne diseases and fecal-oral route diseases | 0.07 |

Mapping inter-connectedness between codes to represent the lived experience of the community (Phase 2 and 3 of the modified FAIR Framework; see Fig 1), revealed a system in which 19 source nodes initiated a system of inter-connected factors with cascading effects reshaping lives and disease trajectories (Table 3):

- The bombing of residential areas displaced populations to overcrowded camps, creating environments conducive to the transmission of communicable diseases such as Hepatitis A. Repeated displacement, compounded by the bombing of camps and surrounding areas, further hindered the establishment and maintenance of safe water and sanitation facilities. These conditions facilitated the spread of waterborne diseases and other infections transmitted via the fecal-oral route, contributing to the progression of severe acute malnutrition in affected children.
- The bombing and destruction of homes, residential buildings, and civilian areas result in deaths, displacement, and significant disruption to family life. Women bear a disproportionate burden, taking on the responsibility for their family’s survival, stability, and serving as moral and cultural anchors for children, often at the expense of their own self-care. This heightened role increases their vulnerability to communicable diseases and maternal health risks. As primary caregivers, women often face challenges in providing effective care, leading to delays in seeking medical attention. Consequently, when individuals finally present at healthcare facilities, their conditions are frequently advanced, critical, or decompensated, making treatment more complex and outcomes less favorable.
- The bombing and destruction of healthcare facilities forced patients to travel longer distances to access care, often under life-threatening conditions. Many patients delayed seeking care due to the risks associated with travel, leading to late-stage presentations requiring referral to tertiary care. These delays compounded the severity of illness, increasing the need for ICU-level care in already resource-constrained environments.
- The prevention of food transport to the population emerged as a critical driver of starvation, forcing individuals to consume toxic materials, such as animal feed, as a last resort. This led to organ toxicity, including hepatotoxicity, and the onset of severe acute malnutrition. Similarly, the restriction of necessary sources of nutrition resulted in insufficient caloric and nutritional intake, further exacerbating SAM. These pathways illustrate how systemic barriers to food access directly influenced disease progression in vulnerable populations, particularly children.
- The prevention of fuel transport to healthcare facilities disrupted essential services, including intensive care unit (ICU) operations. Hospitals were forced to rely on manual ventilation for critically ill patients, leading to inconsistent oxygen delivery, hypoxemia, and hypoxic injury. These disruptions severely compromised patient outcomes, especially for those requiring continuous monitoring and intervention.
- The embargo on medical supplies created critical shortages of laboratory reagents, medications, and equipment necessary for patient monitoring and treatment. This lack of resources resulted in the delayed diagnosis and inadequate management of preventable conditions, escalating disease severity and further burdening the healthcare system.
- The denial of travel for patients outside their area prevented critical referrals and medical evacuations to facilities with higher levels of care. This lack of access to specialized treatment led to the decompensation of patients into multi-organ failure and, ultimately, death.

**Table 3.** Source Nodes (Factors Initiating a System of Inter-Related and Cascading Effects)

| Type of Precipitating Factor | Precipitating Factor |
| --- | --- |
| Bombing & Destruction of People, Infrastructure & Resources | Bombing and destruction of homes, residential buildings, and civilian areas |
|  | Bombing and destruction of camps and areas with displaced civilians |
|  | Bombing and destruction of water treatment plants and sanitation infrastructure |
|  | Bombing and destruction of infrastructure used for water trucking |
|  | Bombing and destruction of aquifers, wells, and desalination plants |
|  | Bombing and destruction of municipal water storage tanks |
|  | Bombing and destruction of Markets and Supply Routes |
|  | Bombing and destruction of roads |
|  | Bombing and destruction of healthcare facilities |
|  | Bombing and destruction of universities and cessation of education |
| Deprivation of Resources | Preventing transport of food to population |
|  | Preventing transport of necessary nutrition sources |
|  | Blockade preventing entry of spare parts and tools |
|  | Shutdown of water lines |
|  | Blockade of transport of essential items (e.g., fuel) |
|  | Blockade of medical supplies |
|  | Blockade and Restrictions on patient travel and medical evacuation |
|  | Blockade and Prevention of healthcare workers to travel within and between Gaza regions |
|  | Blockade of aid; bureaucratic delays in humanitarian aid approval |

Pathways relating to women’s health were unexpectedly significant, with a majority of all pathways in architecture of systems map in some way intersecting with the 13 nodes related to women’s health. In the lived experience of the community, blockades, deprivation, and the bombing and destruction of people, infrastructure and resources, leads to lack of access to supplies necessary for women’s health including hygiene supplies, contraceptives, and prenatal care and nutrition. It also leads to significant behavioral adaptations by communities to survive ongoing death and destruction. These behavioral adaptations to fill the gap of injured, missing, and dead family members results in increasing the burden on women as primary sources of survival for children and families relying on them for obtaining food, water, shelter, and healthcare. It leads to women shouldering a disproportionate burden of responsibility for making decisions about displacement, finding aid, and sustaining the family; in addition to responsibility for the family’s survival and stability, and for being the moral and cultural anchors for children. It places them in roles of surrogate parenthood with a responsibility for the survival and care of children of relatives and neighbors lost to war. These additional burdens deprive communities of societal roles played by women (e.g. as doctors and nurses), leading women to forego self-care to be primary care-givers to others, increasing their susceptibility to infections, worsening their mental health and physical health, delaying treatment until they are in advanced stages of disease. These in turn lead to increased maternal and neonatal morbidity and mortality, and worse outcomes in children relying on them for care.

These findings illustrate that factors brought about by incidents of war interact together to create a system that restricts and shapes choices, movements, and survival strategies to the extent that preventable and self-limiting conditions become fatal. As illustrated in the architecture of systems diagram, war-driven events are the foundational causes leading to a reshaping of lived experiences and resulting in a system optimized to increase morbidity and mortality.

## Discussion

This study demonstrated that anchoring findings in the lived experiences of a population underscores the interconnected factors that shape their health, disease progression, and mortality outcomes.

Many individual pathways in the AoS map are supported by reports that have examined those outcomes. For example, in the AoS map (Fig 1), the community’s experience of restriction of clean water for drinking, hygiene, and healthcare and household daily use was traced to the targeting of water infrastructure by military action. This is validated by reports that document the destruction of water facilities and plants,(30) wells,(31,32) and reservoirs(30,33,34) by airstrikes and other military action,(35) resulting in the reduction of water supply to the population in Gaza.(35) Similarly, injuries, deaths, and displacement from airstrikes, bombings, and attacks on civilian populations and infrastructure has been well documented by other reports.

This study reveals additional pathways that highlight the centrality of women’s health in mediating the effects of military action on civilian populations. The AoS map, comprising 138 nodes and 231 edges, generates 41,444 pathways. Strikingly, only 13 nodes (9%) are directly related to women’s health, yet these nodes are linked to an overwhelming 95% (39,639) of all pathways. Centrality metrics of the AoS map reveal that in Gaza half of the top 10 most crucial links mediating the relationship between the destruction of war and health outcomes relate to the impact of war on women. It should be noted that while women-related nodes speak directly to issues specific to women, other nodes include both men and women (e.g. deaths and injuries that result from bombings). The impact of war on women as described in this study includes: the unshared burden of responsibility borne by women, caused by loss of deaths and injuries of other family members; the loss of women’s participation in critical social roles; women foregoing self-care to be primary care giver for others; worsening mental health for women; and caregivers being unable to provide needed care effectively resulting from impacts on women’s health (see Supplementary File 1, Table A; and Supplementary File 2, List of Pathways). The large number of pathways that intersect with nodes related to women underscores that in the lived experience of the community women’s health plays a pivotal role in mediating the effects of war on population health, particularly on children. The high degree centrality of women-related nodes means that when women are impacted by war-related events, such as bombings and resource destruction, there are downstream effects on health outcomes of children and the community.

The findings in this study reveal a profound and multifaceted impact of war on health outcomes, illustrating how destruction of civilian infrastructure destroys existing systems, engineering a new landscape of survival challenges for communities (Fig 1). It reveals how the effect of military action on civilian populations directly (e.g. bombing residential areas) or indirectly (e.g. destruction of water and sanitation facilities, destruction of crops and greenhouse) imposes a hostile system of influences that forces communities to make choices outside the context of a functioning societal framework. Such impacts of war on civilians fundamentally reshapes the determinants of health by creating circumstances where the very actions taken for survival—like eating substitute foodstuffs in the absence of safe options— become direct pathways to illness and death. This study demonstrates that war in Gaza has not merely interrupted established systems; it has generated a series of forced decisions that actively undermine community health. By removing conventional choices and introducing new, perilous alternatives as survival adaptations, this engineered context escalates vulnerability, establishing an entirely different set of hazards compared to those seen in natural or accidental crises (Ben Taleb et al., 2020).

These findings also emphasize the necessity of contextualizing patient outcomes within the systems producing them. In Gaza, where restrictions limit humanitarian aid(36) and obstruct medical evacuations,(37) patients are prevented from reaching the advanced care they need (Fig 1). Such realities transform the healthcare journey into one fraught with delays, risks, and barriers, significantly altering the usual trajectory of disease progression. For example, the inability to transport critically ill patients out of conflict zones prevented access to life-saving interventions, leading to otherwise avoidable multi-organ failure and death (Fig 1). Understanding patient trajectories within systems shaping them is vital for designing interventions that consider clinical needs while taking into account the obstacles that affect patient access to care.(38)

This study’s findings are shaped by several limitations inherent to research conducted in active conflict zones. The reliance on a modified FAIR Framework, while context-sensitive, required adaptations that may have influenced data comprehensiveness and participant engagement. For instance, condensing multiple sessions into single encounters and substituting patient charts for vignettes might have constrained the depth of narrative enrichment and thematic analysis. Additionally, the destruction of records and infrastructure posed challenges to obtaining consistent data, potentially leading to gaps or biases in the representation of community experiences. The persistent threat to participant safety, including risks associated with documentation, may have further limited the candor and scope of shared insights. Finally, while the study focuses on the lived experiences of affected populations, the findings are specific to the context of North Gaza and may not fully generalize to other conflict settings, where distinct socio-political and infrastructural dynamics could yield different pathways and outcomes. Future research should address these challenges by exploring alternative ways to adapt participatory methodologies to the context of active war, and expanding the scope to include broader geographic and temporal contexts. Additionally, as systems are often dynamic, changing over time, future research can move beyond a cross-sectional snapshot of a system and explore how to represent a changing system as experienced by the effected communities. Representing a dynamic system over time can enable monitoring, analysis, development of projections, and facilitate a dynamic humanitarian response that is sensitive to and anticipatory of evolving needs on the ground.

This study showed that military action on civilian populations engineers systems of worsening morbidity and mortality. More generally, this study demonstrated that incorporating local voices and experiences can provide essential insights into understanding the shaping of community realities by circumstances of war. Clarifying these reshaped systems gives insights into the integrated humanitarian response necessary to mitigate the adversely changed health and life trajectories of affected populations.

## Data Availability

All data that was recorded as part of this study are included in the supplement files (S1 File. Participatory modeling scripts and results S2 File. List of pathways). Detailed transcripts are not available as they were not taken out of the conflict zone for fear of retaliation if electronically associated with the researchers prior to their departure or found on their person or device while transporting them.

## Acknowledgements

Not applicable.

